# Does the Sleep Regularity Questionnaire capture objective sleep–wake regularity? Evidence from wearable and sleep diary data

**DOI:** 10.64898/2026.02.24.26347047

**Authors:** Matthew Driller, Michael E Bodner, Alyssa Fenuta, Shauna Stevenson, Haresh Suppiah

## Abstract

Sleep regularity is an important but under-measured dimension of sleep health. Objective indices from actigraphy or wearables are robust but resource-intensive. The Sleep Regularity Questionnaire (SRQ) offers a brief subjective tool, but its validity against objective and diary-based indices in healthy adults is unclear.

In Part 1, 31 adults wore a smart ring continuously for 21 nights. Device-derived regularity metrics included the Sleep Regularity Index (SRI), interdaily stability (IS), social jetlag (SJL), composite phase deviation (CPD), and the standard deviation of sleep onset and wake time. In Part 2, 52 adults completed a one-week sleep diary, from which variability in sleep timing, total sleep time (TST), SJL and nightly perceived sleep quality were derived. All participants completed the SRQ and Brief Pittsburgh Sleep Quality Index (B-PSQI).

In Part 1, associations between SRQ scores and device-derived SRI, IS, SJL, CPD and timing variability were small (absolute r ≤ 0.36). Higher SRQ Global and Sleep Continuity scores were moderately associated with better B-PSQI global scores (r −0.37 to −0.44). In Part 2, SRQ Global and Circadian Regularity showed small-to-moderate associations with higher diary-rated sleep quality and lower bedtime variability (r ≈ 0.40 and −0.32 to −0.34), while correlations with other diary metrics and B-PSQI were weak (absolute r ≤ 0.25).

The SRQ shows modest convergent validity with diary-based timing variability and perceived sleep quality, but only weak correspondence with smart ring-based sleep regularity indices. It is likely to complement, rather than replace, objective monitoring in healthy adults with relatively regular sleep–wake patterns.

## Introduction

Sleep is increasingly recognised as a multidimensional construct that encompasses duration, quality, timing, efficiency, satisfaction, and regularity (Buysse, 2014). While duration and quality have historically received the most attention, more recent work has highlighted sleep regularity - the consistency of sleep–wake timing across days, as a critical, and previously underappreciated, determinant of health and performance (Phillips et al., 2017; Sletten et al., 2023). Irregular sleep–wake patterns have been linked to impaired cognitive function, elevated cardiometabolic risk, poorer academic and occupational performance, and adverse mental health outcomes (Depner et al., 2019; Huang et al., 2020; Lo et al., 2016).

Sleep regularity is thought to exert its influence largely through circadian alignment (Potter et al., 2016). Stable bed and wake times promote synchrony between internal circadian rhythms and external zeitgebers, such as the light–dark cycle and social schedules (Quante et al., 2019). When sleep timing is inconsistent, misalignment can occur between the central circadian pacemaker and behavioural sleep–wake patterns, leading to autonomic dysregulation, perturbed glucose metabolism, altered appetite and mood regulation, and, over time, increased disease risk (Huang et al., 2020; Wittmann et al., 2006). Importantly, recent studies suggest that individuals with more regular sleep schedules achieve more efficient and consolidated sleep, even when total sleep duration is similar, reinforcing the notion that regularity represents a distinct dimension of sleep health rather than a proxy for duration or quality alone (Phillips et al., 2017). Further, Halson et al. (2022) reported that more regular sleepers tended to obtain longer and more efficient sleep in 203 elite team sport athletes. Similarly, Teece et al. (2024) demonstrated that higher sleep regularity in professional rugby union athletes were associated with longer sleep duration and generally more favourable sleep profiles. These findings suggest that sleep regularity may be a relevant target for sleep optimisation interventions in high-performing individuals.

Objective characterisation of sleep regularity has been facilitated by actigraphy and wearable technologies, which can generate continuous, high-resolution sleep–wake time series across multiple days (Depner et al., 2020; Sadeh, 2011). A range of indices have been proposed. The Sleep Regularity Index (SRI) quantifies the probability of being in the same state (asleep vs awake) at matching clock times on adjacent days intra-individual standard deviations of sleep onset, offset and midsleep capture variability in key timing landmarks; interdaily stability (IS) reflects the strength and consistency of circadian rest–activity rhythms; social jetlag (SJL) captures the discrepancy in sleep timing between workdays and free days; and composite phase deviation (CPD) integrates both nightly variability and mistiming of midsleep (Huang et al., 2020; Phillips et al., 2017; van Someren et al., 1999). Together, these metrics provide a rich description of sleep–wake consistency and its physiological implications.

However, recent work has shown that the way these indices, particularly SRI, are implemented can substantially affect their absolute values and their associations with health outcomes. Czeisler and colleagues systematically compared multiple open-source SRI implementations (including GGIR and sleepreg) across large accelerometry datasets and demonstrated that differences in data source (e.g., raw accelerometry vs device-derived sleep/wake), epoch length, day definition (midnight-to-midnight vs noon-to-noon), and handling of naps and missing data can yield materially different SRI distributions and risk estimates, even when nominally applying the same “SRI” concept (Czeisler et al., 2025). To address this, they proposed the RIRI (Reporting Items for Regularity Indices) checklist, a 14-item reporting framework that specifies, among other elements, the underlying data type, preprocessing steps, epoch length, day anchoring, definitions of sleep and wake, software or package used, and how missingness and outliers are handled. This framework emphasises that regularity indices are not plug-and-play, and that transparent, standardised reporting is essential for comparability across studies and for translation to clinical and public health contexts.

Despite the increasing sophistication of objective regularity metrics, their deployment in many research and applied settings is constrained by cost, logistics, and analytic expertise. Actigraphy and advanced wearable devices require high compliance, careful data cleaning, and access to specialised software, and the resulting indices can be difficult to interpret outside specialist groups. This has driven growing interest in brief, scalable, and psychometrically sound subjective measures of sleep regularity that can complement or, where necessary, substitute for device-based assessment, provided they are properly validated against well-characterised objective and diary-derived indices and reported in line with frameworks such as RIRI.

The Sleep Regularity Questionnaire (SRQ) was developed as a brief, 10-item instrument to assess perceived consistency in sleep timing and duration (Dzierzewski et al., 2021). Exploratory and confirmatory factor analyses indicated that, of the original 10 items, a subset of six consistently loaded onto two reliable subscales: circadian regularity (bedtime, wake time, rise time, and sleep duration regularity) and sleep continuity regularity (regularity of nocturnal awakenings and time awake at night). The remaining four items (e.g., napping, feeling refreshed) showed weaker or inconsistent loadings and were excluded, yielding a 6-item version with improved psychometric properties. To date, however, no study has directly compared the SRQ with well-specified objective or diary-based indices of sleep regularity in healthy adults while explicitly situating those indices within the methodological guidance provided by Czeisler et al. (2025) and the RIRI checklist. As a result, it remains unclear to what extent the SRQ captures the same underlying regularity construct quantified by device-derived sleep regularity metrics.

The present study therefore aimed to validate the SRQ using a two-part design. In Part 1, we compared SRQ scores with objective regularity indices derived from 21 nights of continuous monitoring with a smart ring. SRI, IS, CPD, social jetlag, and timing variability were calculated from a binary sleep–wake series using a custom implementation of the original SRI formula that adheres to key RIRI-recommended reporting elements (e.g., explicit specification of epoch length, day definition, and handling of naps and missing data). In Part 2, we examined associations between SRQ scores, and diary-derived variability metrics computed from 7 nights of daily sleep diary entries in a larger sample, providing a parallel validation using self-reported timing. In both parts, the Brief Pittsburgh Sleep Quality Index (B-PSQI) was administered to assess convergent validity with global sleep quality. We hypothesised that SRQ scores, particularly the circadian and sleep continuity subscales, would be positively associated with higher SRI and IS, and negatively associated with intra-individual variability, social jetlag, CPD, and diary-based variability in sleep timing. We also expected SRQ scores to be inversely related to PSQI scores, reflecting the broader association between greater regularity and better subjective sleep quality.

## Materials & Methods

### Study Design

This study employed a two-part, observational validation design. Part 1 assessed the convergent validity of the SRQ against objective indices of sleep regularity derived from wearable monitoring, while Part 2 assessed the validity of the SRQ against diary-derived variability metrics. All participants completed the SRQ and B-PSQI. The study was conducted in accordance with the Declaration of Helsinki and approved by the institutional Human Research Ethics Committee (HEC22369). Written informed consent was obtained from all participants. Sample sizes for both parts of the study were determined pragmatically, balancing statistical precision with the burden of multi-night monitoring and diary completion. In Part 1, 31 adults each contributed 21 consecutive nights of wearable-derived sleep–wake data (>650 person-nights). In Part 2, 52 adults completed a 7-night sleep diary (∼364 person-nights). The study was designed as an exploratory validation of the SRQ against complementary objective and diary-based indices, rather than to detect very small effect sizes. The achieved sample sizes provide adequate power to detect small-to-moderate correlations (r ≈ 0.30–0.40) between SRQ scores and regularity indices, and are appropriate for this stage of construct validation in healthy adults.

### Participants

Participants were healthy adults aged 20–65 years recruited via university mailing lists and word of mouth. Exclusion criteria included: diagnosed sleep, psychiatric, or neurological disorders; current use of sleep medication; night or rotating shift work in the previous 3 months; and travel across two or more time zones in the previous 4 weeks. A total of 35 participants were originally recruited for Part 1 (later reduced to 31 due to missing data), providing 21 nights of wearable data (18 males [58%], 13 females [42%]; mean age = 33.5 ± 9.7 y). In Part 2, 52 healthy adults participated (27 males [52%], 25 females [48%]; mean age = 21.9 ± 3.2 y). In both Part’s 1 and 2 of the study, participants were told to sleep as they normally would, keeping their usual habits and patterns. Across both Parts 1 and 2, participants were instructed to maintain their usual sleep–wake schedules and behaviours throughout the monitoring period. They were informed that they would complete subjective sleep questionnaires, including an assessment of perceived sleep regularity, at the end of the study, but were not advised or incentivised to modify their sleep patterns; thus, any systematic reactivity toward increased regularity is considered unlikely. The individuals in Part 1 and 2 of the study were not the same, therefore the entire study included 83 individual participants.

### Sleep Regularity Questionnaire

Sleep regularity was assessed using a shortened version of the Sleep Regularity Questionnaire (SRQ). From the original 10 items, only six were retained based on prior factor-analytic work demonstrating that these items load onto a two-factor structure representing (i) circadian regularity and (ii) sleep continuity regularity (Dzierzewski et al., 2021). The four items relating to consistent bedtimes, wake times, rise times, and sleep duration formed the *circadian regularity* subscale, reflecting the stability of sleep–wake timing across days. The two items relating to the number and duration of nocturnal awakenings formed the *sleep continuity regularity* subscale, reflecting the stability of sleep maintenance across nights. Each item was rated on a 5-point Likert scale (0 = *not at all* to 4 = *very much*), with higher scores indicating greater regularity (see Table 1). Subscale scores were calculated by summing their respective items, and a global SRQ score was derived by summing across all six items. The SRQ was completed before midday, but at least one hour after waking up (mean time 11:13am).

**Table 1.**
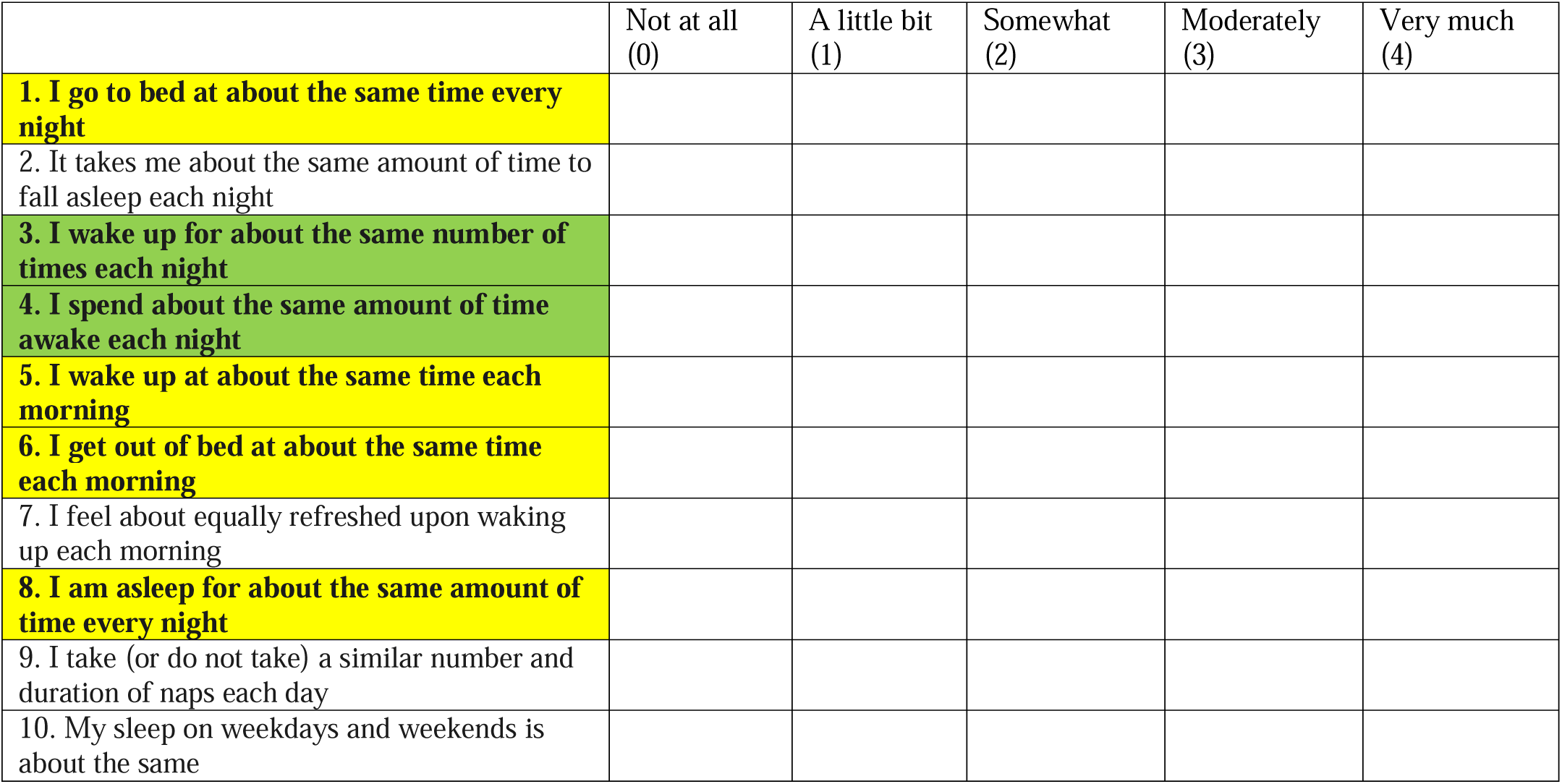
The Sleep Regularity Questionnaire (Dzierzewski et al., 2021), including the original 10-item tool, the reduced 6-items in bold (used in the current study). The Circadian Regularity subscale is highlighted in yellow, and the Sleep Continuity subscale in green.

### Brief Pittsburgh Sleep Quality Index (B-PSQI)

Subjective sleep quality was assessed using the Brief Pittsburgh Sleep Quality Index (B-PSQI), a shortened version of the original 19-item PSQI (Sancho-Domingo et al., 2021). The B-PSQI retains six core items that capture the major dimensions of sleep: (i) subjective sleep quality, (ii) sleep latency, (iii) sleep duration, (iv) habitual sleep efficiency, (v) sleep disturbances, and (vi) daytime dysfunction. Item responses are combined into five component scores (0–3 each), which are then summed to generate a global B-PSQI score ranging from 0 to 15, with higher totals reflecting poorer sleep quality. Consistent with the original PSQI, a global score >5 is commonly used to indicate clinically relevant sleep disturbance. The brief version has demonstrated comparable psychometric properties to the full PSQI while reducing participant burden (Sancho-Domingo et al., 2021).

### Objective Sleep Monitoring

In Part 1 of the current study, participants wore a smart ring (Ultrahuman Ring AIR, Ultrahuman Healthcare Pvt Ltd, Bangalore, India) continuously for 21 consecutive nights. The device integrates triaxial accelerometry, photoplethysmography, and skin temperature sensors. Validation of the smart ring against a criterion device (Somfit, single-channel EEG) demonstrated excellent agreement for sleep onset (r = 0.9998, ICC = 0.9997) and sleep offset (r = 0.985, ICC = 0.974), and good agreement for total sleep time (r = 0.889, ICC = 0.844, see Supplementary Files, Supplementary Table 1 for comparison data).

For the present analyses, we exported device-derived sleep data as 5-minute epochs labelled as awake, light, deep or REM sleep. Epochs outside the automatically detected nocturnal sleep period (bedtime–wake time) were recorded but, by design, all such epochs were treated as wake in the regularity analyses. From these data, we derived a binary sleep–wake series (1 = asleep for any of light/deep/REM; 0 = awake) and computed several regularity metrics: the Sleep Regularity Index (SRI), interdaily stability (IS), social jetlag, composite phase deviation (CPD), and within-person variability in bed and wake times. Full details of these regularity metrics are provided below.

### Derivation of the Sleep Regularity Index (SRI)

We calculated the Sleep Regularity Index (SRI) to quantify the day-to-day consistency of sleep–wake timing, following the original formulation by Phillips et al. (2017) and the detailed description in the RIRI (Reporting Items for Regularity Indices) statement (Czeisler et al., 2025). SRI was calculated using a custom Python implementation of the original Phillips et al. (2017) equation, following the noon-to-noon, −100 to +100 convention used in GGIR, rather than using the R packages GGIR or sleepreg directly, because only device-derived 5-minute sleep/wake epochs (and not raw accelerometry) were available.

For each participant and each recording day, we constructed a 24-hour time series from 12:00 (noon) to 11:55 the following day in 5-minute epochs (288 epochs per 24 h). Within the nocturnal sleep window identified by the Ultrahuman algorithm, epochs labelled as light, deep or REM sleep were coded as sleep (1), and epochs labelled as awake were coded as wake (0). All epochs outside the nocturnal sleep window (i.e., daytime and evening periods not classified as sleep by the device) were set to wake (0). Daytime naps were not present in this population and therefore did not contribute to the SRI calculation. The SRI was calculated as:

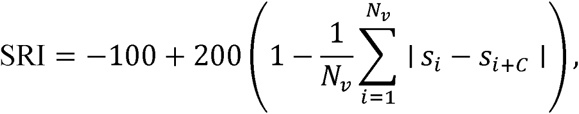

where *s_i_* denotes the sleep–wake state at epoch *i* (e.g., 1 = asleep, 0 = awake), and *C* is the number of epochs in 24 h (here, *C* = 288). For each valid pair of epochs separated by exactly 24 h, we compared the state at times *i* and *i* +*C*, and *Nv* is the number of such valid epoch pairs. This yields an SRI score on a – 100to +100scale, where +100reflects regularity (approximately half the pairs matched and half mismatched), and – 100reflects perfectly regular sleep–wake timing (state always identical 24 h apart), 0 reflects no net perfectly reversed regularity (state always opposite 24 h apart).

### Interdaily Stability

Interdaily stability (van Someren et al., 1999) was calculated as a non-parametric rest–activity regularity metric, adapted here for binary sleep–wake state. Using the same 24-hour, 5-minute epoch series described above, each participant’s data were arranged into days (rows) by 5-minute epochs (columns). The sleep–wake state at epoch *i*is denoted. *x_i_*(0 = wake, 1 = sleep), *p*is the number of epochs per day (*p* =288), and *N* is the total number of epochs included. The overall mean state is *x̄*, and the mean state at each clock-time bin *h* is *x̄_h_*. Interdaily stability was then:

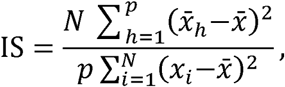

such that values closer to 1 indicate a more stable, clock-time–anchored pattern of sleep and wake across days.

### Social jetlag (SJL)

Social jetlag was defined as the absolute difference between mid-sleep on “free” nights versus “work” nights (Wittmann et al., 2006). For each night we identified sleep onset (first epoch coded as sleep) and final wake time (end of the last sleep epoch), and calculated midsleep as the midpoint of this interval. “Work nights” were defined as nights with wake-up on Monday–Friday, and “free nights” as those with wake-up on Saturday or Sunday. For each participant, circular mean midsleep was computed separately for work and free nights, and SJL was expressed in hours as the absolute circular difference between these two means.

### Composite phase deviation (CPD)

Composite phase deviation was used to characterise night-to-night irregularity in sleep timing (Fischer et al., 2016). For each participant, midsleep times for all nights were expressed on a circular 24-h scale. For each night *i*, two circular differences in hours were calculated: the difference between midsleep on night *i* and the participant’s circular mean midsleep (μ), and the difference between midsleep on night *i* and midsleep on the previous night (*x*_*i* − 1_). These were computed as shortest circular distances (i.e., wrapped to the −12to +12h range). The nightly CPD was then

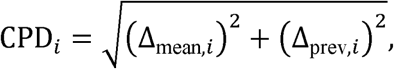

where Δ*_mean,i_* is the circular difference between midsleep on night *i* and μ, and Δ*_prev,i_* is the circular difference between midsleep on night *i* and night i − 1. Composite phase deviation was summarised as the mean CPD*_i_* across nights (from night 2 onward), with larger values indicating more mistimed and irregular sleep timing.

### Variability in bed and wake times

As additional markers of behavioural regularity, we computed circular mean and circular standard deviation (SD) of bedtime and wake time across the recording for each participant. Bedtime was defined as the first epoch of the nocturnal sleep window, and wake time as the end of the last sleep epoch. Times were converted to minutes after midnight on a circular 24-hour scale, and circular SD (in minutes) was used as an index of within-person variability in bed and wake timing.

### Quality control and adherence

In line with recommendations from the RIRI statement, we required at least five consecutive 24-hour intervals with valid SRI comparisons for inclusion in the primary analyses (Czeisler et al., 2025). For a given participant, epoch pairs with missing data at either time point were excluded from the SRI denominator (*Nv* ), but days were retained if at least 50% of potential epoch pairs for that 24-h lag were available. All participants included in the final analyses for Part 1 met these criteria and contributed ≥5 days of valid SRI/IS data, however, four participants were excluded on the basis of missingness or wear time, giving a total of 31 participants.

### Sleep diary and derived metrics

In Part 2 of the current study, participants completed a 7-day sleep diary in which they entered their habitual bedtime (clock time), waketime (clock time), perceived sleep quality (0–10 numerical rating), and their total sleep time. From these nightly entries we derived a set of night-level variables and then computed participant-level aggregates.

For each night, bedtime and waketime were taken directly from the reported clock times in the diary. Mid-sleep was calculated as the midpoint between bedtime and waketime for that night. To avoid cross-midnight artefacts, all clock times were first converted to minutes since a “day anchor” of 04:00; times earlier than 04:00 were temporarily shifted by +24 h, mid-sleep was computed on this unwrapped scale, and the resulting values were then mapped back to the conventional 24-h clock.

For each participant we summarised the 7 diary nights into week-level metrics. Mean bedtime and its standard deviation (SD) were calculated on the unwrapped minute scale to characterise both the average bedtime and its variability across the week. The same approach was used to derive mean waketime and SD waketime. Mean mid-sleep and SD mid-sleep were computed from the nightly mid-sleep values described above. In addition, we calculated mean TST and SD TST to describe average sleep duration and its night-to-night variability. Mean perceived sleep quality was obtained by averaging the nightly SQ ratings across the week.

Social jet lag was operationalised as the absolute difference between the participant’s mean mid-sleep on free days (weekend nights with wake-up on Saturday or Sunday) and mean mid-sleep on workdays (weeknights with wake-up Monday to Friday). When a diary week did not include both workdays and free days, social jet lag was set to missing for that participant.

### Statistical Analysis

For Part 1, analyses focused on the convergent validity of the Sleep Regularity Questionnaire (SRQ) with objective indices of sleep–wake regularity derived from the smart ring. All analyses were conducted on the full sample of participants with valid SRQ scores and ring-derived regularity metrics (n = 31); no outliers were removed, consistent with a priori decisions and to preserve the ecological range of variability. Continuous variables were inspected for distributional characteristics using histograms and summary statistics. Descriptive statistics (mean, standard deviation, minimum, maximum) were calculated for all objective regularity metrics including; Sleep Regularity Index (SRI), interdaily stability (IS), social jetlag (SJL), composite phase deviation (CPD), and the standard deviation of bedtime and waketime, as well as for SRQ scores and B-PSQI global scores.

The primary convergent validity analyses used Pearson correlations to quantify associations between each SRQ score (Circadian Regularity, Sleep Continuity Regularity, and Global) and each objective regularity metric (SRI, IS, SJL, CPD, bedtime variability, waketime variability). Correlation coefficients (r) and p-values are reported for each SRQ–metric pair. Because this was an exploratory validation study with a modest sample size, correlations are interpreted primarily in terms of effect size and direction; p-values are provided for completeness but not used as the sole determinant of importance. In secondary analyses, simple linear regression models were fitted with each objective metric as the outcome, SRQ Global as the main predictor, and age and sex included as covariates. These models were used to verify whether demographic factors altered the strength or significance of the SRQ–regularity associations. In all cases, inclusion of age and sex did not materially change the pattern or magnitude of the associations, so the primary results are presented as unadjusted correlations for simplicity, with the adjusted models briefly summarised in the text. All analyses were performed in Python (pandas, SciPy, statsmodels), with two-sided α set at 0.05.

In Part 2, all analyses were conducted in Python (pandas, numpy, SciPy, statsmodels, scikit-learn). Two-sided tests were used throughout and α was set at 0.05 unless noted otherwise. Sleep-diary variables supplied in HH:MM format were converted to minutes for analysis to ensure consistent scaling across outcomes. Diary outcomes were computed at the participant level and included weekly standard deviations (SD) of bedtime, waketime, midsleep, and total sleep time (TST); weekly means of TST and perceived sleep quality (/10); and social jetlag, defined as the absolute difference in midsleep between workdays and free days (minutes).

Criterion validity was assessed by estimating Spearman rank correlations between each SRQ score (SRQ Global, Circadian Regularity, Sleep Continuity) and each diary outcome (bedtime SD, waketime SD, midsleep SD, social jetlag, TST SD, mean TST, perceived sleep quality). The false discovery rate was controlled using the Benjamini–Hochberg procedure across the full set of SRQ × outcome tests, and both raw and FDR-adjusted p-values are reported. To evaluate whether associations with timing variability were driven by typical schedule, a sensitivity analysis was performed using partial correlations that controlled for the corresponding mean timing (e.g., SRQ vs bedtime SD adjusted for mean bedtime).

As an applied secondary analysis, the ability of SRQ scores to identify individuals with high sleep-timing variability was examined, with “high variability” defined a priori as the top tertile of midsleep SD. Discrimination was summarised using receiver-operating characteristic area under the curve (ROC-AUC) with 5-fold stratified cross-validation to limit optimism. No additional feature selection or model tuning was undertaken, given the sample size and the screening intent of this analysis.

Analyses were conducted on complete cases per outcome (i.e., participants were included in each correlation or model if they had the requisite SRQ score and diary outcome). Summary statistics (means, SDs, quartiles, and ranges) are reported for all SRQ scores and diary outcomes. Effect sizes (Spearman’s r) are presented alongside 95% confidence intervals and p-values; for the classification analysis, cross-validated AUC is reported with its fold-to-fold variability.

In additional exploratory analyses, we examined associations between SRQ scores and the B-PSQI global score in the diary sample (Part 2). Spearman rank correlations were estimated between each SRQ scale (Circadian Regularity, Sleep Continuity, SRQ Global) and B-PSQI global. Given the small number of tests (three correlations), p-values were not adjusted for multiple comparisons; these analyses were intended to position SRQ scores relative to an established questionnaire-based index of overall sleep quality rather than as primary outcomes.

## Results

### Part 1

All summary statistics for Part 1 are included in Supplementary Files (Supplementary Table 2). Objective regularity metrics indicated a generally regular sleep–wake pattern in this cohort. Mean SRI was 80.64 ± 5.61 (range 66.78–90.48) on the −100 to +100 scale. IS was likewise high (0.76 ± 0.07, range 0.53–0.91), consistent with a relatively stable, clock-time–anchored rhythm. Night-to-night variability in timing was moderate: the standard deviation of bedtime was 59.0 ± 25.7 min (range 25.2–135.7 min), and the standard deviation of waketime was 58.1 ± 24.4 min (range 20.6–125.8 min). Social jetlag averaged 0.90 ± 0.88 h (range 0.09–3.92 h), and composite phase deviation (CPD), reflecting combined mistiming and night-to-night variability of midsleep, was 1.10 ± 0.50 h (range 0.47–2.89 h). Overall, participants were objectively quite regular, with relatively small weekday–weekend shifts.

SRQ scores also suggested moderate to high perceived regularity. The Circadian Regularity subscale had a mean of 10.29 ± 2.90 (range 2–15), indicating that most participants reported fairly consistent bed and wake times and sleep duration. The Sleep Continuity Regularity subscale averaged 4.84 ± 2.21 (range 1–8), reflecting greater variability in perceived regularity of awakenings and time awake at night. The SRQ Global score was 15.13 ± 4.11 (range 4–23). B-PSQI global scores averaged 3.87 ± 2.54 (range 1–12), consistent with generally good subjective sleep quality in this sample.

As hypothesised, SRQ scores showed positive but generally small associations with the device-derived Sleep Regularity Index (SRI) and interdaily stability (IS). Across the three SRQ scales, correlations with SRI and IS were all weak (r ≲ 0.20, all p > 0.35; Table 2), and scatterplots of SRQ Global versus SRI and IS showed minimal linear trends (Figures 1 and 2).

**Figure 1.**
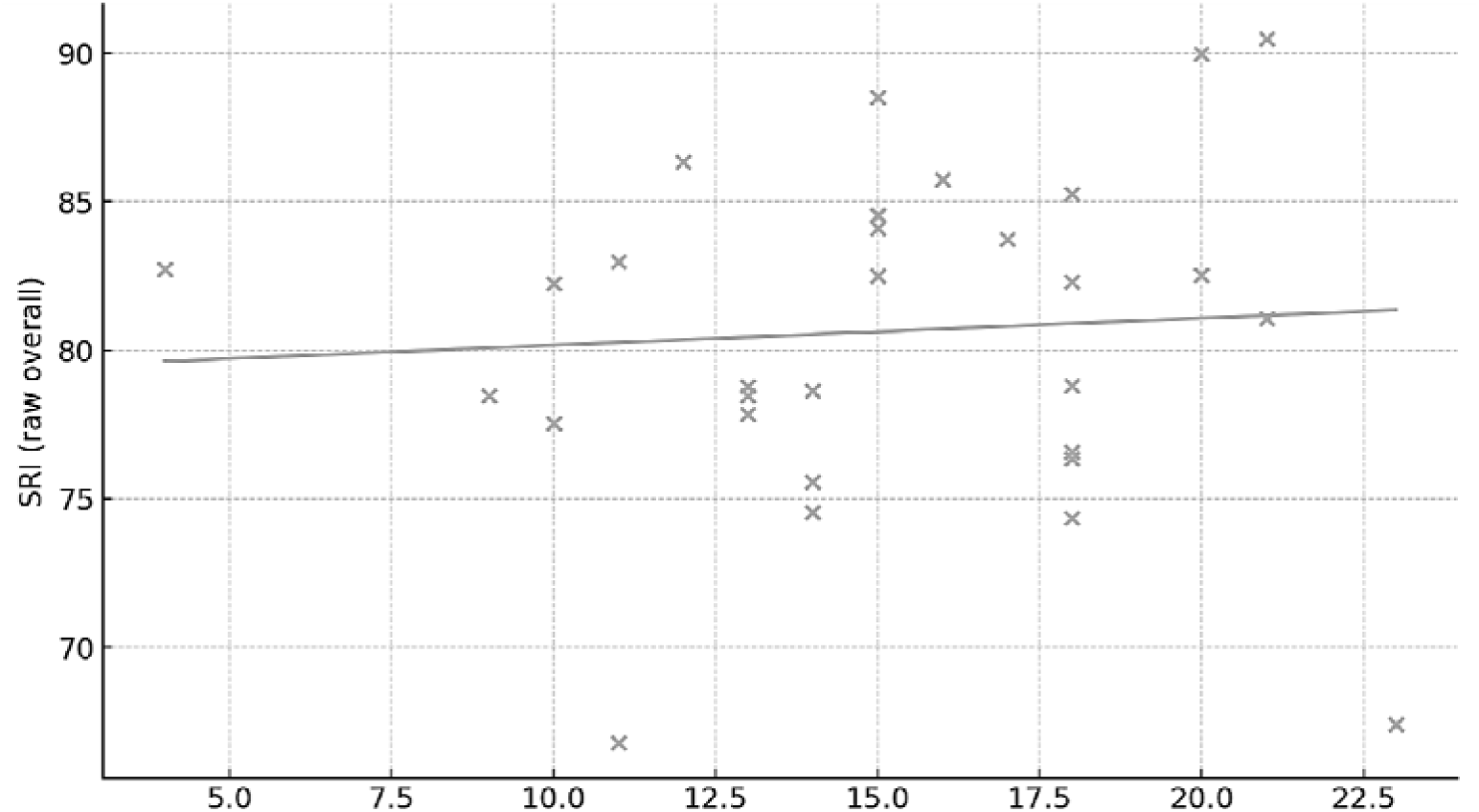
Association between SRQ Global score and device-derived Sleep Regularity Index (SRI) in adults wearing a smart ring (n = 31). Each point represents one participant’s SRQ Global score and ring-derived SRI averaged across the monitoring period.

**Figure 2.**
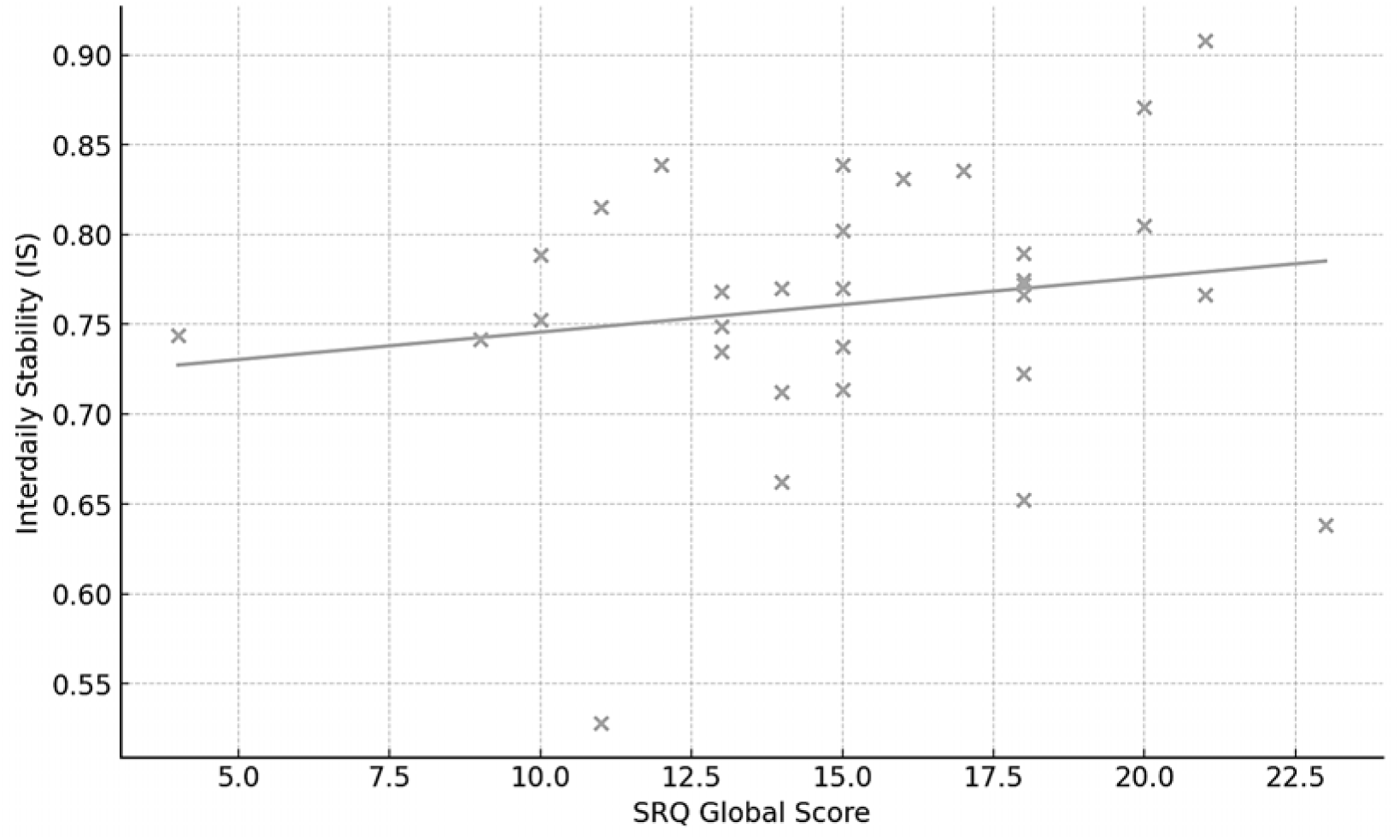
Association between SRQ Global score and device-derived interdaily stability (IS) in adults wearing a smart ring (n = 31). Each point represents one participant’s SRQ Global score and IS computed from the actigraphy-derived sleep–wake series.

**Table 2.** Pearson correlations (r) between SRQ scores and objective sleep–wake regularity metrics derived from the smart ring (n = 31). Pearson r values include 95% confidence intervals in parentheses. p is the two-sided unadjusted p-value for each correlation.

| Objective regularity metric | Circadian<br>Regularity<br>r | p | Sleep Continuity<br>Regularity<br>r | p | SRQ Global<br>r | p |
| --- | --- | --- | --- | --- | --- | --- |
| Sleep Regularity Index (SRI) | 0.012<br>(-0.344, 0.365) | 0.949 | 0.108<br>(-0.256, 0.445) | 0.562 | 0.067<br>(-0.295, 0.411) | 0.722 |
| Interdaily stability (IS) | 0.123<br>(-0.242, 0.458) | 0.509 | 0.154<br>(-0.212, 0.482) | 0.408 | 0.170<br>(-0.197, 0.494) | 0.362 |
| Social Jetlag (SJL) | 0.357<br>(0.003, 0.631) | 0.049 | 0.181<br>(-0.185, 0.503) | 0.330 | 0.349<br>(-0.006, 0.626) | 0.054 |
| Composite Phase Deviation (CPD) | 0.273<br>(-0.090, 0.572) | 0.137 | 0.127<br>(-0.238, 0.461) | 0.495 | 0.261<br>(-0.103, 0.563) | 0.156 |
| Bedtime variability | 0.281<br>(-0.081, 0.578) | 0.125 | 0.062<br>(-0.299, 0.408) | 0.739 | 0.232<br>(-0.133, 0.542) | 0.209 |
| Waketime variability | 0.079<br>(-0.284, 0.421) | 0.674 | 0.036<br>(-0.322, 0.386) | 0.846 | 0.075<br>(-0.287, 0.418) | 0.689 |

Correlations between SRQ scores and other timing-variability metrics (social jetlag, CPD, bedtime SD, waketime SD) were also modest (Table 2). The largest effects were observed for social jetlag: Circadian Regularity was moderately correlated with social jetlag (r = 0.36, p = 0.049), with a similar trend for SRQ Global (r = 0.33, p = 0.07). Associations with CPD and bedtime/waketime variability were small-to-moderate and positive (r ≈ 0.2–0.3), but none reached conventional significance. Overall, these findings suggest that, in this healthy adult, non shift-working cohort, self-reported regularity on the SRQ aligns only weakly with device-derived 24-h regularity metrics.

Exploratory linear regression models including age and sex as covariates did not materially change the associations between SRQ Global and SRI or IS; SRQ effects remained small and non-significant, and neither age nor sex was independently related to these objective regularity metrics. Primary inferences are therefore based on the unadjusted correlations in Table 2.

For context, subjective sleep quality showed clearer relationships with both objective regularity and the SRQ. Higher B-PSQI global scores (worse sleep quality) were moderately associated with lower SRI and IS (r = −0.36 and −0.42, respectively), whereas correlations with social jetlag, CPD, and timing variability were small (|r| ≤ 0.12). SRQ scores were inversely related to B-PSQI, particularly for Sleep Continuity Regularity (r = −0.44) and SRQ Global (r = −0.37), indicating that individuals who perceived more regular sleep, especially in terms of continuity, also reported better overall sleep quality, despite only weak correspondence with detailed device-derived regularity metrics in this sample.

### Part 2

All summary statistics for Part 2 are included in Supplementary Files (Supplementary Table 3). SRQ scores showed their clearest associations with perceived sleep quality and bedtime variability (Figure 3, Table 4). SRQ Global and Circadian Regularity were moderately correlated with higher mean perceived sleep quality (both r ≈ 0.41, FDR-adjusted p = 0.051; Figure 4, Table 4). Correlations with bedtime variability were in the expected negative direction (more regular SRQ scores associated with lower bedtime SD; r ≈ −0.32 to −0.34) but did not remain significant after FDR correction (Table 4). Associations with variability in waketime, midsleep, social jetlag, TST variability, and mean TST were small and non-significant across all SRQ scales (|r| ≤ 0.25, FDR-adjusted p ≥ 0.14; Figure 3, Table 4).

**Figure 3.**
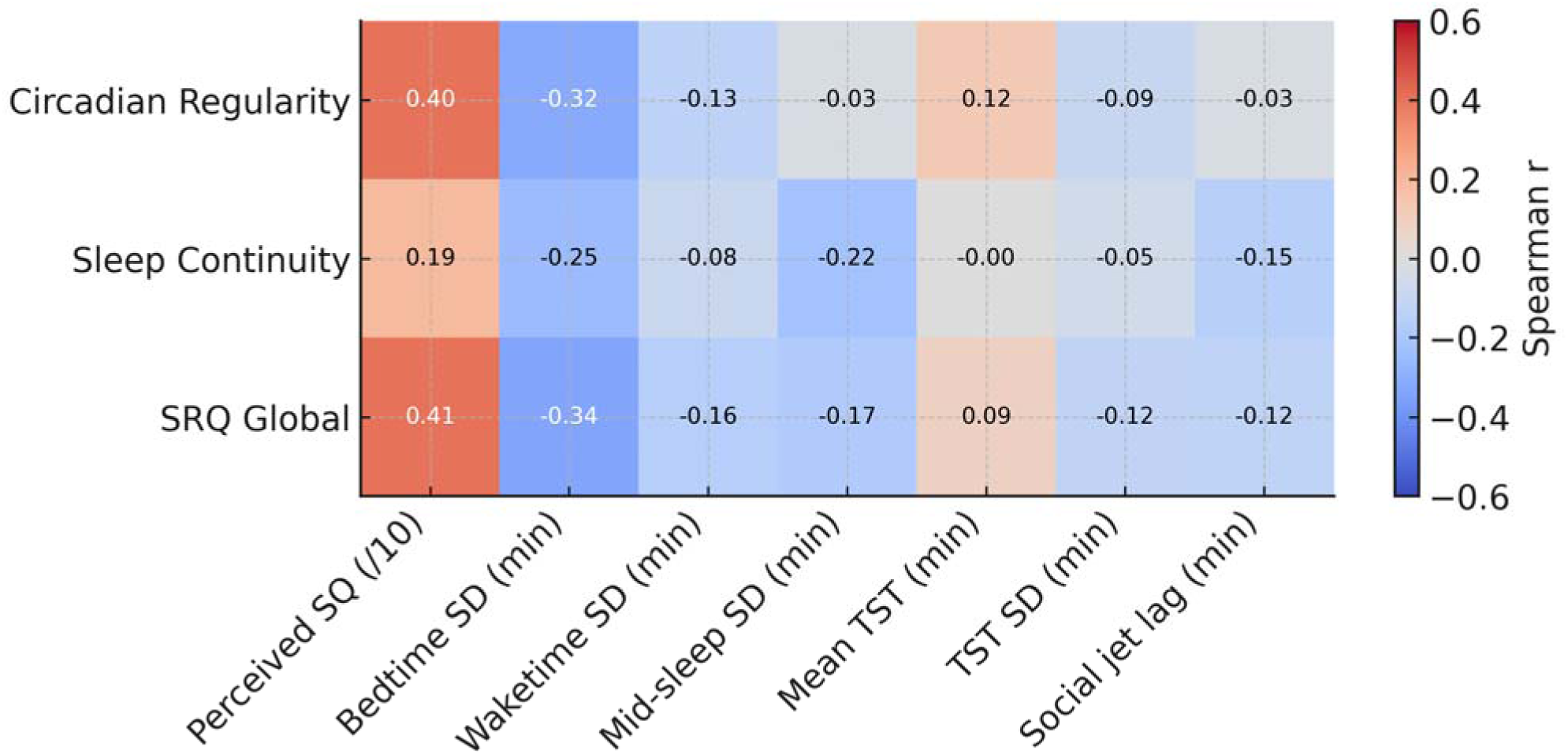
Spearman correlations between SRQ scores and diary-derived sleep outcomes (n = 52). Rows show the three SRQ scales (Circadian Regularity, Sleep Continuity, and SRQ Global score), and columns show participant-level diary outcomes (perceived sleep quality, variability [SD] in bedtime, waketime, midsleep and total sleep time (TST), mean TST, and social jetlag).

**Figure 4.**
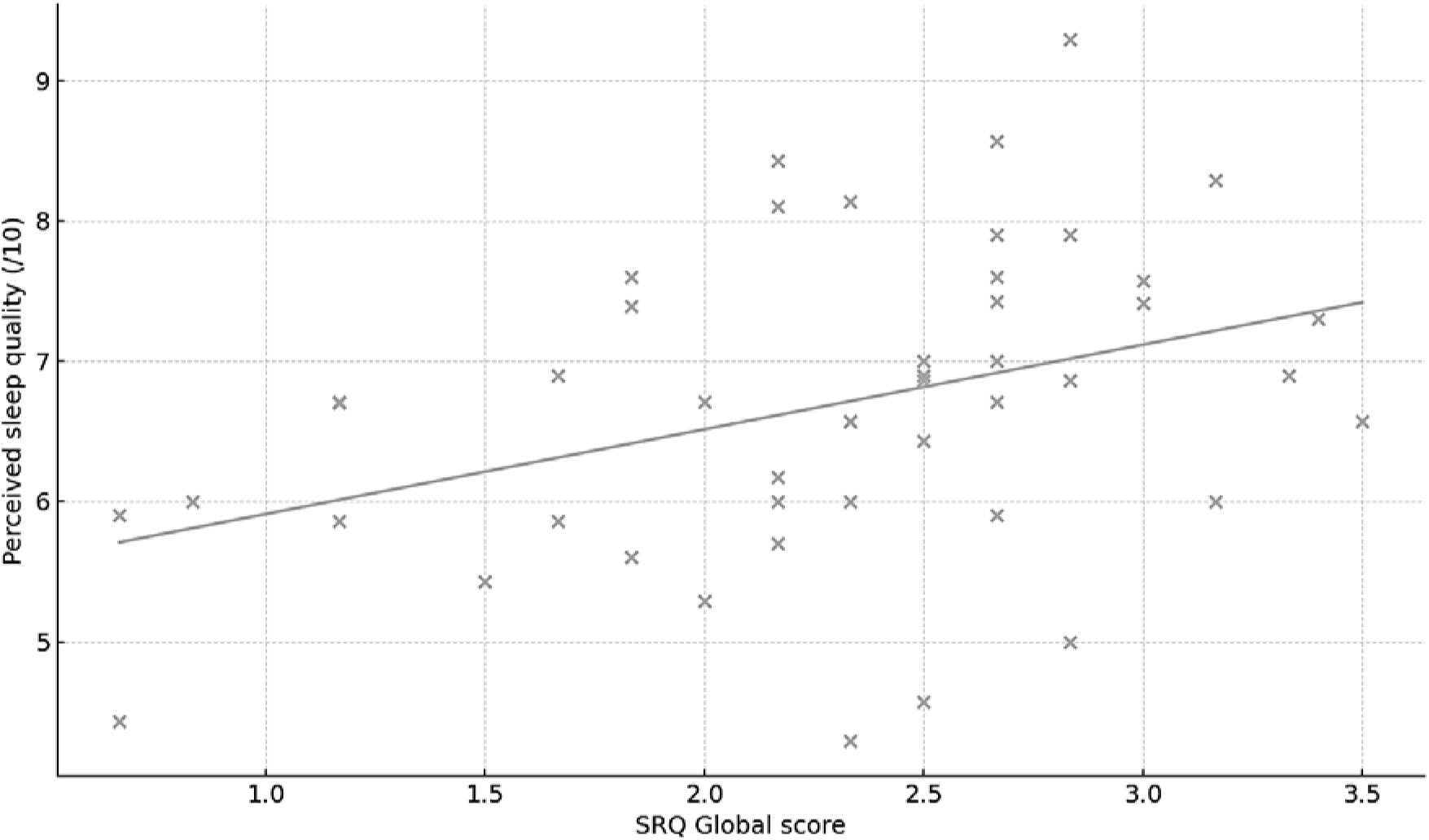
Association between SRQ Global score and perceived sleep quality in the sleep-diary sample (n = 52). Each point represents one participant’s SRQ Global score (x-axis) and mean perceived sleep quality over 7 nights (/10; y-axis).

**Table 4.** Spearman correlations (r) between SRQ scores and diary-derived sleep outcomes. Values are Spearman r with 95% confidence intervals in parentheses. p is the two-sided unadjusted p-value for each correlation. p (FDR) is the corresponding p-value adjusted for multiple testing using the Benjamini–Hochberg false discovery rate procedure across all SRQ × outcome combinations (3 SRQ scores × 7 diary outcomes).

| Diary outcome | Circadian<br>Regularity<br>r | p | p<br>(FDR) | Sleep<br>Continuity<br>r | p | p<br>(FDR) | SRQ Global<br>r | p | p<br>(FDR) |
| --- | --- | --- | --- | --- | --- | --- | --- | --- | --- |
| Perceived sleep quality (/10) | 0.404<br>(0.132, 0.619) | 0.005 | 0.051 | 0.191<br>(-0.102, 0.454) | 0.198 | 0.685 | 0.407<br>(0.135, 0.621) | 0.005 | 0.051 |
| Bedtime variability | -0.303<br>(-0.553, -0.003) | 0.048 | 0.254 | -0.274<br>(-0.531, 0.028) | 0.075 | 0.316 | -0.352<br>(-0.590, -0.058) | 0.021 | 0.144 |
| Waketime variability | -0.064<br>(-0.351, 0.234) | 0.676 | 0.878 | -0.045<br>(-0.334, 0.252) | 0.768 | 0.896 | -0.101<br>(-0.383, 0.198) | 0.509 | 0.801 |
| Mid-sleep variability | -0.005<br>(-0.305, 0.296) | 0.977 | 0.986 | -0.188<br>(-0.462, 0.120) | 0.228 | 0.685 | -0.150<br>(-0.431, 0.158) | 0.338 | 0.789 |
| Mean Total Sleep Time | 0.153<br>(-0.140, 0.422) | 0.303 | 0.789 | 0.011<br>(-0.277, 0.297) | 0.943 | 0.986 | 0.108<br>(-0.185, 0.383) | 0.469 | 0.801 |
| Total Sleep Time variability | -0.062<br>(-0.349, 0.236) | 0.688 | 0.878 | -0.057<br>(-0.345, 0.241) | 0.711 | 0.878 | -0.107<br>(-0.389, 0.192) | 0.483 | 0.801 |
| Social jetlag (SJL) | -0.003<br>(-0.306, 0.301) | 0.986 | 0.986 | -0.128<br>(-0.415, 0.183) | 0.421 | 0.801 | -0.099<br>(-0.391, 0.212) | 0.534 | 0.801 |

In a sensitivity analysis controlling for mean bedtime, the association between SRQ Global and bedtime variability changed little, suggesting that the link between perceived regularity and bedtime variability is not solely driven by typical bedtime. In an applied secondary analysis, SRQ scores showed limited ability to discriminate individuals in the highest tertile of midsleep variability, with cross-validated ROC–AUC of 0.52 (SD 0.23 across five folds), close to chance, indicating modest screening utility for identifying highly irregular sleepers in this sample.

Finally, we examined convergent validity with the Brief Pittsburgh Sleep Quality Index (B-PSQI). In the diary sample, B-PSQI scores averaged 4.5 ± 2.6 (range 1–14), with approximately one-third of participants scoring ≥5. SRQ scores were weakly and non-significantly associated with B-PSQI, with small negative correlations for Circadian Regularity (r = −0.15, p = 0.33), Sleep Continuity (r = −0.16, p = 0.29) and SRQ Global (r = −0.14, p = 0.36), indicating that participants who perceived more regular sleep tended to report slightly better overall sleep quality, but the effects were modest. As expected, higher (worse) B-PSQI scores were moderately associated with lower mean diary-rated sleep quality (/10; r = −0.39, p = 0.007) and shorter mean total sleep time (r = −0.54, p < 0.001), whereas associations with variability in bedtimes, waketimes, midsleep, social jetlag and TST were small (|r| ≤ 0.22, all p ≥ 0.17).

## Discussion

This study examined the validity of the Sleep Regularity Questionnaire (SRQ) against multiple indices of sleep–wake regularity derived from a wearable device (Part 1) and a one-week sleep diary (Part 2) in healthy adults. Across 21 nights of ring-based monitoring, participants in the device subsample showed relatively high objective regularity (mean SRI ∼81, IS ∼0.76, social jetlag < 1 h), and SRQ scores indicated moderate–high perceived regularity. The central finding from Part 1 is that correlations between SRQ scores and device-derived regularity metrics (SRI, IS, SJL, CPD, and variability in sleep and wake times) were small (absolute r ≤ 0.36), whereas SRQ scores, particularly the Sleep Continuity subscale and SRQ Global, showed moderate associations with subjective sleep quality indexed by the B-PSQI. In Part 2, SRQ Global and Circadian Regularity showed small-to-moderate associations with higher diary-rated perceived sleep quality and modest associations with lower bedtime variability, while relationships with other diary-based regularity metrics and B-PSQI were weak.

These results extend the original SRQ work, which demonstrated a two-factor structure (circadian regularity and sleep continuity regularity) and robust associations with insomnia severity and global sleep quality in a large online adult sample, while emphasising that the SRQ captures a construct related to, but distinct from, global sleep disturbance (Dzierzewski et al., 2021). Subsequent translation and validation studies have replicated the two-factor structure and shown that lower SRQ scores are associated with poorer subjective sleep quality and greater insomnia symptoms, as well as higher stress, anxiety and depression (Yan et al., 2024; Zhang & Qin, 2023). Other work has linked lower sleep regularity scores to greater emotion-regulation difficulties and poorer subjective cognitive functioning, even after accounting for insomnia severity and global sleep health (Nielson et al., 2025; Perez et al., 2022). Together with our findings, this pattern suggests that individuals who endorse higher regularity on the SRQ tend to report better sleep quality and better emotional or cognitive functioning, but they do not necessarily exhibit higher regularity metrics as determined by wearable devices. Interestingly, higher social jetlag was modestly associated with higher SRQ Circadian and Global scores in the device subsample (Part 1). This likely reflects that SJL quantifies differences in timing between workdays and free days, whereas the SRQ does not explicitly distinguish between these contexts. Individuals who maintain a consistent pattern on workdays and a similarly consistent (but shifted) pattern on free days may therefore experience their sleep timing as ‘regular’ and endorse high SRQ scores, despite exhibiting measurable social jetlag. Given the restricted range of social jetlag in our study sample, these associations should be interpreted cautiously.

The weak correspondence between SRQ scores and regularity metrics from a wearable device in the current study is notable given accumulating evidence that actigraphy-based regularity metrics relate to important health outcomes, including cardiometabolic risk, adiposity, depressive symptoms and mortality (Depner et al., 2019; Huang et al., 2020; Phillips et al., 2017; Zhang & Qin, 2023). Several non-mutually exclusive explanations are plausible. First, the constructs are not identical. The SRQ focuses on the perceived consistency of bedtimes, wake times and consolidated nocturnal sleep windows over weeks to months, whereas SRI and IS quantify the probability of being in the same state (asleep vs awake) at specific clock times. It is therefore possible, in general, for individuals to perceive their main sleep episode as regular while still exhibiting irregular state patterns at the edges of the night or during daytime naps; however, in the present sample daytime naps were not observed, so discrepancies are more likely to arise from behaviour at the margins of nocturnal sleep. Second, our sample showed restricted variability in both SRI and SRQ scores, which limits the observable correlations. Third, SRQ responses require long-term recall (7 to 21 days in our study) and compress multiple behaviours into a small number of Likert ratings, introducing recall and anchoring biases that may attenuate associations with high-resolution actigraphy.

More broadly, the modest convergence we observed is consistent with a large literature documenting only small-to-moderate correspondence between subjective and objective sleep measures. Habitual self-reported sleep quality and duration often show weak or inconsistent associations with polysomnography- or actigraphy-derived metrics such as sleep efficiency and total sleep time in community and clinical samples (Cudney et al., 2022; Kaplan et al., 2017). Recent work similarly indicates that objective indicators of sleep continuity and architecture explain only a modest proportion of variance in night-to-night subjective sleep ratings (Pierson-Bartel & Ujma, 2024). A recent scoping review of physiological markers of sleep quality and a broader synthesis of sleep-discrepancy research conclude that subjective and objective measures capture overlapping but distinct aspects of sleep and cannot be treated as interchangeable proxies (Walton et al., 2025). Within this context, it is perhaps unsurprising that a brief retrospective questionnaire such as the SRQ aligns more strongly with other subjective indices (e.g., B-PSQI, diary-rated quality) than with high-resolution regularity metrics derived from wearables.

By contrast, agreement with diary-based indices was clearer, particularly for perceived sleep quality. In Part 2, higher SRQ Global and Circadian Regularity scores were associated with better diary-rated sleep quality, with correlations around 0.40, and showed small negative correlations with bedtime variability. Although the bedtime associations did not remain significant after FDR correction and associations with variability in waketime, midsleep, social jetlag and TST were small (absolute r ≤ 0.25), the overall pattern is consistent with the SRQ capturing a subjective sense of stable sleep timing that aligns more strongly with how people feel about their sleep than with fine-grained day-to-day timing variability. This mirrors work showing that sleep regularity indices from actigraphy relate to mood, cardiometabolic risk and daytime functioning independent of average sleep duration (Depner et al., 2019; Huang et al., 2020; Phillips et al., 2017; Zhang & Qin, 2023), but also underscores that device-based metrics and brief questionnaires are not interchangeable.

Several limitations should be acknowledged. The sample was relatively small and comprised healthy, predominantly non-shift-working adults from a single country, which may limit generalisability to older, clinical or highly irregular populations such as shift workers. Participants in both parts of the study showed relatively regular sleep schedules, with high SRI and modest social jetlag, which likely constrained the range of regularity captured by both the SRQ and device metrics and may have attenuated correlations. Existing SRQ studies in larger and more heterogeneous samples have shown that SRQ scores vary meaningfully across demographic and health strata and relate to mental health and cognitive outcomes (Dzierzewski et al., 2021; Nielson et al., 2025; Yan et al., 2024; Zhang & Qin, 2023), suggesting that the questionnaire can capture a broad spectrum of regularity; however, convergent validity with objective regularity metrics in shift workers or clinical populations has yet to be established. The diary period in Part 2 was limited to one week, which may not fully characterise habitual variability, and objective regularity metrics were derived from a single wearable device and proprietary sleep–wake classification; although our validation work suggests good agreement with a research-grade monitor, residual measurement error cannot be excluded. Finally, the cross-sectional design precludes conclusions about sensitivity to change; it remains unclear how responsive the SRQ is to interventions targeting sleep regularity.

Practically, these findings suggest that the SRQ is best viewed as a brief index of perceived sleep timing regularity and continuity that complements, rather than replaces, device-based metrics of sleep regularity. In large-scale or resource-limited settings where actigraphy is not feasible, the SRQ may offer a pragmatic way to capture an aspect of sleep regularity that is meaningfully related to sleep quality and, to a lesser extent, diary-based timing variability. For work specifically targeting SRI-like constructs that quantify 24-hour state regularity, multi-day device-based monitoring remains essential. Future research should test the SRQ in populations with greater irregularity (for example, shift workers, individuals with circadian rhythm sleep–wake disorders, or athletes during congested competition schedules), examine its responsiveness to interventions, and integrate SRQ, diary and device measures within multivariate models to identify the most prognostic aspects of "sleep regularity" for health and performance.

## Data Availability

All data produced in the present study are available upon reasonable request to the authors

## Acknowledgements

The authors would like to acknowledge the team at Ultrahuman Healthcare Ltd – Aditi Bhattacharya, Nihav Dhawale, Ved Asudani, and Kanika Gupta, for providing the smart rings for this study, and for their assistance with data extraction.

## Supplementary Files

### Reliability of Ultrahuman Ring Device

Sleep was measured concurrently using the Somfit (criterion device) and Ultrahuman Ring Air (test device) across 20 nights in 4 participants (2 male, 2 female; mean ± SD age = 29 ± 6 years) to ascertain key sleep metrics used in the current study (sleep onset, sleep offset, and total sleep time).

- Somfit: Criterion data were acquired using the Somfit system, which records single-channel EEG (Fp1–Fp2), electro-oculography (Fp1–Fpz, Fp2–Fpz), electromyography (Fp1–Fp2), pulse oximetry, head position, snoring sound, movement, and ambient light. Somfit has been validated against full polysomnography (Miller et al., 2022; Roach et al., 2025).
- Ultrahuman Ring Air: A commercial wearable ring integrating photoplethysmography (PPG), accelerometry, and skin temperature sensors. Proprietary algorithms are used to derive sleep timing and duration.

Sleep onset, sleep offset, and total sleep time were extracted for each night. Agreement between devices was assessed using Pearson correlations, intraclass correlation coefficients (ICC; two-way mixed, absolute agreement), mean difference (bias), typical error (TE), and coefficient of variation (CV%).

### Results

Across 20 paired nights, agreement between devices was excellent for sleep onset (r = 0.9998, ICC = 0.9997) and sleep offset (r = 0.985, ICC = 0.974), and good for total sleep time (r = 0.889, ICC = 0.844). Average differences between devices were small (–1.4 to +3.6 min), with typical errors of 8–15 min.

**Supplementary Table 1.** Validity statistics (20 nights) between the Ultrahuman Ring and Somfit (criterion).

| Measure | Somfit<br>Mean ± SD | Ultrahuman<br>Mean ± SD | Pearson's<br>$r$ | ICC | Mean<br>Diff<br>(min) | TE (min) | CV (%) |
| --- | --- | --- | --- | --- | --- | --- | --- |
| Sleep Onset | 22:55 ± 1:12 h | 22:55 ± 1:12 h | 0.9998 | 0.9997 | -1.4 | 8.3 | 0.9 |
| Sleep Offset | 07:17 ± 0:59 h | 07:21 ± 1:02 h | 0.985 | 0.974 | +3.6 | 8.0 | 1.8 |
| Total Sleep | 7:44 ± 0:45 h | 7:45 ± 0:44 h | 0.889 | 0.844 | +1.0 | 14.9 | 3.2 |

**Supplementary Table 2.** Mean data from Part 1 of the study, including objective sleep metrics collected by the smart ring over 21 days (n=31) and SRQ scores.

| **Metric** | **Mean ± SD** | **Range** | **Units / notes** |
| --- | --- | --- | --- |
| Mean bedtime | 21:57 ± 4.1 h | 00:02 – 23:58 | Clock time; within-participant mean bedtime (SD reflects between-participant dispersion of mean bedtimes) |
| Mean wake time | 06:57 ± 0.9 h | 04:28 – 08:32 | Clock time; within-participant mean wake time |
| Sleep Regularity Index (SRI) | 80.64 ± 5.61 | 66.78 – 90.48 | –100 to +100; higher values indicate more regular timing |
| Interdaily stability (IS) | 0.76 ± 0.07 | 0.53 – 0.91 | Dimensionless; higher values indicate more stable, clock-time–anchored rhythms |
| Bedtime variability (SD of bedtime) | 59.0 ± 25.7 min | 25.2 – 135.7 | Minutes; within-participant SD of nightly bedtime |
| Waketime variability (SD of waketime) | 58.1 ± 24.4 min | 20.6 – 125.8 | Minutes; within-participant SD of nightly waketime |
| Social jetlag (SJL) | 0.90 ± 0.88 h | 0.09 – 3.92 | Hours; absolute difference in midsleep between workdays and free days |
| Composite phase deviation (CPD) | 1.10 ± 0.50 h | 0.47 – 2.89 | Hours; combined mistiming and night-to-night variability of midsleep |
| SRQ Circadian Regularity subscale | 10.29 ± 2.90 | 2 – 15 | Sum of 4 items (0–16); higher scores indicate more regular bed and wake timing |
| SRQ Sleep Continuity Regularity subscale | 4.84 ± 2.21 | 1 – 8 | Sum of 2 items (0–8); higher scores indicate more regular nocturnal awakenings and time awake |
| SRQ Global score | 15.13 ± 4.11 | 4 – 23 | Sum of 6 items (0–24); higher scores indicate more regular perceived sleep timing and continuity overall |

**Supplementary Table 3.** Mean data from Part 2 of the study, including subjective sleep diary metrics collected over one week (n=52) and SRQ scores.

| **Metric** | **Mean ± SD** | **Range** | **Units / notes** |
| --- | --- | --- | --- |
| Mean bedtime (diary-derived) | 23:44 ± 1.37 h | 00:06 – 23:57 | Clock time; within-participant mean bedtime |
| Mean wake time (diary-derived) | 07:43 ± 1.97 h | 03:40 – 20:49 | Clock time; within-participant mean wake time |
| Perceived sleep quality | 6.69 ± 1.11 | 4.29 – 9.29 | Diary-rated sleep quality (0–10; higher = better) |
| Mean total sleep time (TST) | 7.53 ± 1.18 h | 4.42 – 10.00 | Hours; within-participant mean nightly TST |
| Bedtime variability (SD of bedtime) | 114.9 ± 138.8 min | 10 – 676 | Minutes; within-participant SD of nightly bedtime |
| Waketime variability (SD of waketime) | 87.6 ± 99.0 min | 22 – 669 | Minutes; within-participant SD of nightly waketime |
| Mid-sleep variability (SD of midsleep) | 104.5 ± 150.9 min | 9 – 725 | Minutes; within-participant SD of nightly midsleep |
| TST variability (SD of TST) | 75.6 ± 43.2 min | 20 – 262 | Minutes; within-participant SD of nightly TST |
| Social jetlag | 1.23 ± 1.06 h | 0.05 – 4.90 | Hours; absolute difference in midsleep between workdays and free days |
| SRQ Circadian Regularity subscale (sum) | 8.64 ± 2.84 | 0 – 14 | Sum of 4 items (0–16); higher scores indicate more regular bed and wake timing |
| SRQ Sleep Continuity Regularity subscale | 5.02 ± 2.09 | 0 – 8 | Sum of 2 items (0–8); higher scores indicate more regular awakenings and time awake at night |
| SRQ Global score | 13.66 ± 4.08 | 4 – 21 | Sum of 6 items (0–24); higher scores indicate more regular perceived sleep timing and continuity overall |

## Notes

### Competing Interest Statement

The authors have declared no competing interest.

### Funding Statement

This study did not receive any funding

### Author Declarations

Human Research Ethics Committee at La Trobe University gave ethical approval for this work

